# Seroprevalence of Human *Brucella* Antibodies and associated Among Patients Seeking Medical Attention at Community Hospitals in Selected Districts of Western Province in Zambia

**DOI:** 10.1101/2024.07.25.24311011

**Authors:** Armand Mayindu Mambote, John Bwalya Muma, Mary Mubiana, Steward Mudenda, Victor Daka, Melai Mubanga, Flavien Nsoni Bumbangi, Chanda Chitalu, Ruth Lindizyani Mfune

## Abstract

**Introduction:** Brucellosis is a neglected zoonotic disease that affects humans and animals and can lead to severe illness in humans and financial losses for households that rear livestock. The study aimed to investigate the seroprevalence of human *Brucella* antibodies and associated risk factors among patients seeking medical attention at community hospitals.

**Methods:** A cross-sectional seroepidemiological study was conducted from 21^st^ April 2021 to 21^st^ April 2024 among patients seeking medical attention at community hospitals in selected districts of Western province in Zambia. 225 blood samples were collected from consenting participants. Sera were separated and analysed for anti-*Brucella* antibodies using the Rose Bengal Test (RBT) and Competitive enzyme-linked immunosorbent assay (c-ELISA) in parallel. A questionnaire was administered to obtain epidemiological data related to exposure to the *Brucella* pathogen. The data obtained were coded and entered in the Micro-Soft Excel 2013® and analysed using STATA version 15®.

**Results:** 197 sera samples were found acceptable for testing and analysis for this study, out of these, the seroprevalence of *Brucella* antibodies was 18.3% (n=36, 95% CI=0.13-0.24) and 4.57% (n=9, 95% CI=-0.68-0.28) on RBT and c-ELISA respectively.Among the risk factors considered, the number of animals was statistically significantly associated with *Brucella* seropositivity (OR 6.49, 95% CI=1.10-38.13, p-value = 0.039).

**Conclusions:** *Brucella* antibodies are prevalent among patients attending health facilities in the Western province of Zambia. The number of animals were significantly associated with the *Brucella* seropositivity.

## Introduction

*Brucellosis* is an infectious zoonotic disease commonly known as “undulant fever”, “Mediterranean fever”, “gastric remittent fever”, or “Malta fever’’ in humans ^1,2,3,4^. The disease affects humans, wild animals and domestic livestock ^5^. Of the 12 currently known *Brucella* species, only *B. melitensis*, *B. abortus*, *B. suis,* and, on rare occasions, *B. canis* are responsible for human infections^2^. Humans can become infected by consuming unpasteurised dairy products or through direct contact with secretions from infected animals^6^. High-risk occupational groups that are mostly affected include veterinarians, laboratory workers, abattoir workers, slaughterhouse workers, livestock caretakers, and farmers ^7^. Symptoms include fever, headaches, physical weakness, sweats, and back pain^52^. Humans are incapacitated by the condition, which causes significant debility and a loss of active workdays ^9^. The main clinical signs in cattle include abortion, reproductive failure and decreased milk production and 10% of infertility in animals is attributed to *brucellosis*^10^. Abortion-related brucellosis is estimated to be between 30% and 80% in dairy herds under traditional management ^11^. The economic losses caused by these problems are enormous for farmers and at the national level in countries where the disease is endemic ^12,13^.

Brucellosis is among the top seven neglected zoonotic diseases^5^ that continues to have a significant impact globally^14,15^. The World Health Organization (WHO) reports that more than 500,000 new human cases are reported every year globally ^5^. Brucellosis is a significant public health issue identified as an occupational disease^16^. Studies on human *brucellosis* have reported varying seroprevalences in Egypt 31.3% ^17^; Nigeria 24.1% ^18^; Cameroon 5.6% ^19^; Kenya 5.7% and 31.8%^20^; Uganda 17% ^21^ and Tanzania 1.41% ^22^. Due to diverse clinical presentation diverse clinical presentation but on the inadequate diagnostic tools for Brucella in most primary healthcare facilities and also considering that Brucellosis is not part of the routine check in the PHC facilities., only 50% to 60% of cases are detected and recorded^23,4^.

In Zambia, brucellosis is endemic among traditional cattle keepers in the Southern and Western provinces of Zambia, where the practice of raw milk consumption is high^24^. Brucellosis studies have focused more on animals than humans ^24,25,26^. A few studies of human brucellosis have estimated seroprevalences of 5.0%^23,27^ on occupationally exposed individuals (farmers, abattoir workers). However, studies are scarce on clinical cases in hospitals where patients seek routine health services ^27^. Furthermore, laboratory testing for human *brucellosis* is not routinely done in hospitals.

Therefore, this study aimed to fill the existing knowledge gap by determining the seroprevalence of human *Brucella* antibodies and associated risk factors among patients seeking medical attention at community hospitals in selected districts of Western province in Zambia.

## Materials and methods

### Study area and design

A cross-sectional study was conducted from 21^st^ April 2021 to 21^st^ April 2024 among patients seeking medical attention at community hospitals and health facilities in Senanga, Limulunga and Mongu districts of Western Province in Zambia. The province was purposively selected because it is one of the major livestock-producing provinces where brucellosis has been reported ^24^. The three districts were selected because they are home to the central plain where farmers take cattle for grazing. Blood samples were collected from symptomatic patients who visited the health facilities following their consent. A questionnaire was administered to the participants to collect epidemiological information.

### Sample size and Sampling strategy

The sample size was estimated using the Ausvetepitool software (http://epitools.ausvet.com.au/) assuming an expected prevalence of 20% ^31^, a desired absolute precision of 5% and a confidence level of 95%. A minimum sample size of 225 participants was therefore required. The sample size was distributed in the three districts according to the weight index (human population) for each district (Table 1).

**Table 1:** Sample size of humans weighted per district.

| District | Weighting index (Human Population) | Number of person to be sampled |
| --- | --- | --- |
| Mongu | 197816 | 120 |
| Senanga | 12040 | 68 |
| Mongu | 61102 | 37 |
| All areas | **370958** | **225** |

These individuals were grouped into categories depending on their level of daily activities that could lead to direct contact with suspected *Brucella*-infected animals or the use of infected animal products. Three health facilities were selected in Senanga District namely Ngundi, Sikumbi and Lui Wanya Health Centres, while two were included from Mongu (Lealui Mini Hospital and Sefula Rural Health Centre) and Limulunga Districts (Limulumga Mini Hospital and Ikwichi Rural Health Centre) respectively. Sampling was stratified according to districts weighted using the human population as proxy weighting value for the persons to be sampled.

### Inclusion criteria

Patients in the selected hospitals during the study period who presented any of the following signs and symptoms: intermittent or persistent fever, headache, weakness, profuse sweating, chills, arthralgia, weight loss and joint pain fever, with a negative result for malaria were included in the study.

### Exclusion criteria

Patients with associated confirmed disease diagnoses other than Brucellosis were excluded from the study.

### Sampling and Sample Collection

Epidemiological data was collected from consenting participants using a structured questionnaire which was adopted from a similar study by Mubanga et al 2021. The questionnaire consisted of four parts: (i) sociodemographic characteristics of the study participants; (ii) types of slaughtering activities; (iii) work hygiene-related factors (i.e., wearing personal protective equipment, contact with blood or faeces, and presence of skin wound); and (iv) other potential risk factors (cattle breeding, and consumption of raw beef, by-products and milk). The questionnaire was pretested in three similar districts before the commencement of the study and minor corrections were made accordingly. From each participant, four (4) ml of blood was collected by a clinical officer and stored in sterile plain tubes at +4 °C for24-48 hours. The serum samples were separated using a portable field centrifuge (TOMy Digital MX-300, Japan) and stored in labelled cryovial tubes at −20 ◦C until transportation to the University of Zambia, School of Veterinary Medicine for laboratory analysis.

### Laboratory analysis

Serum samples were screened for *Brucella* antibodies using the Rose Bengal Test (RBT, ID.Vet, innovative Diagnostics, France) followed by a confirmation on c-ELISA (SVANOVIR^R^ *Brucella* –Ab c-ELISA, Boehringer Ingelheim Svanova, Sweden) according to the manufacturer’s guidelines and reagent kit manual. A sample was considered positive if any visible sign of agglutination was observed.

### Data analysis

The epidemiological data obtained was coded and entered in Microsoft Excel 2016®, cleaned, exported and analysed using STATA version 17 (Stata Corp., College Station, TX, USA) for Windows. Categorical data were expressed in percentage, and seroprevalence was calculated by dividing the number of positive sera samples by the total samples examined. The odds ratio, 95% confidence interval, and Fisher’s exact tests were computed to see the degree of association of the risk factors with *Brucella* seropositivity. Using the cut-off of P.I.≥ 30% and P.I.≥ 50% for c-ELISA respectively, the independent effects of categorical risk factors on anti-*Brucella* spp. Seropositivity was assessed using Fisher’s exact test. Variables with a *p*-value ≤ 0.25 from the univariable analysis were selected and included in the multivariable logistic model. The multivariable model was built using a backward selection strategy, using a *p*-value of <0.05 of the likelihood ratio test as inclusion criteria. The model fit was assessed using the Hosmer Lemeshow test, *lroc* and *lsens* procedures in Stata for logistic models.

## Results

### Socio-demographic characteristics of study participants

The study had more female 110 (55.8%) than male 87 (44.2%) participants. The mean age of the participants was 36 years, ranging from 10 to 81 years. More than half, 107 (54.31%) of the participants were married. Most participants, 109 (55.33%) had achieved a primary level of education, and 107 (54.31%) were married. Most participants, 134 (68.02%), were unemployed. Senanga District had more participants, 93 (48.23), than Mongu and Limulunga districts (Table 2).

**Table 2:** Socio-demographic variables of study participants.

| Variables | Categories | N = 197 | Per cent | 95% CI | P-value |
| --- | --- | --- | --- | --- | --- |
| **Gender** | Male | 110 | 55.84 | 0.20-2.50 | 0.592 |
|  | Female | 87 | 44.16 | 0.37-0.51 |  |
| **Age** | 10-20 | 49 | 24.97 | 0.28-1.47 | 0.811 |
|  | 21-35 | 52 | 26.40 | 1.19-1.46 |  |
|  | 36-60 | 79 | 40.10 | 0.11-0.05 |  |
|  | >60 | 17 | 8.63 | 0.15-0.10 |  |
| **Level of education** | None | 15 | 7.61 | 0.33-2.37 | 0.924 |
|  | Primary | 109 | 55.33 | 0.13-0.11 |  |
|  | Secondary | 62 | 33.50 | 0.13-0.12 |  |
|  | Tertiary | 7 | 3.55 | 0.27-0.14 |  |
| **Marital status** | Single | 73 | 37.06 | 0.64-4.20 | 0.627 |
|  | Married | 107 | 54.31 | 0.04-0.09 |  |
|  | Divorced | 9 | 4.57 | 0.20-0.11 |  |
|  | Widowed | 8 | 4.06 | 0.08-0.25 |  |
| **Occupation** | Abattoir worker | 1 | 0.51 | 0.49-2.45 | 0.225 |
|  | Health worker | 3 | 1.52 | 0.56-0.85 |  |
|  | Livestock farmer | 25 | 12.69 | 0.13-0.05 |  |
|  | Student | 26 | 13.19 | 0.04-0.20 |  |
|  | Other | 8 | 4.06 | 0.25-0.10 |  |
|  | Unemployed | 134 | 68.02 | 0.53-0.38 |  |
| **District** | Mongu | 66 | 33.50 | 0.08-0.06 | 0.975 |
|  | Limulunga | 36 | 18.27 | 0.10-0.84 |  |
|  | Senanga | 93 | 48.23 | 0.12-0.78 |  |

### Seroprevalence of human *Brucella* antibodies

From the 197 sera samples that were acceptable for testing and analysis, the estimated seroprevalence of *Brucella* antibodies among the patients attending the community hospitals in Western Province was 18.3% (n=36, 95% CI=0.13-0.24) on RBT and 4.57% (n=9, 95% CI=- 0.6814-0.2807) on c-ELISA (p-=0.412). The seroprevalence was higher in Senanga and Mongu (2.54%) than Limulunga district (Table 3).

**Table 3:** Distribution of *Brucella* antibody seropositivity per study district.

| DISTRICT | Number tested | RBT | C-ELISA |
| --- | --- | --- | --- |
|  |  | Seropositive participants (%) | Seropositive participants (%) |
| **Mongu** | 66 | 10 (5.08) | 4 (2.03) |
| **Senanga** | 93 | 19 (9.64) | 4 (2.03) |
| **Limulunga** | 38 | 7 (3.55) | 1 (0.51) |
| **Total** | **197** | **36 (18.27%)** | **9 (4.57%)** |

### Knowledge and attitudes of participants regarding Brucellosis

Most of the participants 114 (57.87%) had obtained their information on brucellosis from veterinary officers, while only a few, 13 (6.60%), were aware that brucellosis can affect humans. Most participants, 132 (67.01%), were ignorant about the mode of transmission to humans while only 16 (8.12%) stated the symptoms of brucellosis as shown in Table 4.

**Table 4:** Knowledge and attitude of participants about Brucellosis.

| Variables | Categories | N =197 | Per cent | 95% C.I | P-value |
| --- | --- | --- | --- | --- | --- |
| **Information about Brucellosis** | Yes | 114 | 57.87 | 0.09-0.47 | 0.8 |
|  | No | 83 | 42.13 | 0.08-5.03 |  |
| **Source of information**<br>**N=114** | Veterinary officer | 53 | 46.49 | 0.98-5.67 | 0.734 |
|  | Neighbour | 12 | 10.53 | 0.76-2.56 |  |
|  | Health worker | 39 | 34.21 | 0.59-1.81 |  |
|  | Media | 4 | 3.50 | 0.08-3.46 |  |
|  | Patients with Brucellosis | 6 | 5.26 | 0.34-7.23 |  |
| **Can humans be affected?** | Yes | 13 | 6.60 | 0.54-6.86 | 0.663 |
|  | No | 67 | 34.01 | 0.87-4.89 |  |
|  | I do not know | 117 | 59.39 | 0.47-4.11 |  |
| **Mode of transmission** | I do not know | 132 | 67.01 | 0.58-8.14 | 0.167 |
|  | Contact with an infected animal | 25 | 12.69 | 0.87-9.76 |  |
|  | Eating undercooked meat and drinking raw milk | 40 | 20.3 | 0.91-1.84 |  |
| **Symptoms of Brucellosis** | Yes | 16 | 8.12 | 0.61-2.38 | 0.004 |
|  | No | 181 | 91.88 | 0.62-3.24 |  |

### Hygienic and protection practices of study participants regarding Brucellosis

A high proportion of the participants drank raw milk 27 (13.71%) and consumed undercooked meat 170 (86.29%). Most participants kept animals 114 (57.87%), and among these about 40 (48.19%) had more than ten animals, while 17 (20.48%) had their animals vaccinated against brucellosis (Table 5).

**Table 5:** Hygienic and vaccination characteristics of study participants about Brucellosis.

| **Variables** | **Categories** | **N (197)** | **Per cent** | **95% C.I** | **P-value** |
| --- | --- | --- | --- | --- | --- |
| **Drinking raw milk and eating undercooked meat** | No | 27 | 13.71 | 0.57-5.84 | 0.647 |
|  | Yes | 170 | 86.29 | 0.46-4.65 |  |
| **Keeping animal** | No | 114 | 57.87 | 33.99-39.16 | 0.818 |
|  | Yes | 83 | 42.13 | 0.68-6.87 |  |
| **Type of animal owned (N=83)** | Only goat | 14 | 16.87 | 0.76-9.75 | 0.308 |
|  | Only sheep | 19 | 22.89 | 0.25-0.456 |  |
|  | Only pigs | 8 | 9.64 | 9.87-24.45 |  |
|  | Cattle | 25 | 30.12 | 1.92-27.82 |  |
|  | Dogs | 9 | 10.84 | 2.92-26.75 |  |
|  | Other | 8 | 9.64 | 8.89-28.76 |  |
| **Number of animals**<br>**(N=83)** | One animal | 5 | 6.02 | 8.96-22.37 | **0.049** |
|  | 1-10 animals | 38 | 45.79 | 3.14-42.17 |  |
|  | >10 animals | 40 | 48.19 | 4.67-15.67 |  |
| **Vaccinated animal**<br>**(N=83)** | Yes | 17 | 20.48 | 7.896-21.76 | 0.421 |
|  | No | 66 | 79.52 | 5.87-19.65 |  |

### Clinical signs and symptoms of the study participants

The main reason for seeking medical attention by most participants was fever 161 (81.72%) with a p-value of 0.004 followed by malaise, 17 (8.63%). Participants who had the onset of symptoms from 2 to 6 days, 161 (81.72%), were the most likely to attend the health facilities (Table 6).

**Table 6:** Clinical symptoms of study participants in the study area (Mongu, Limulunga, Senanga)

| Variables | Categories | N (197) | Per cent | CI | P-value |
| --- | --- | --- | --- | --- | --- |
| Symptoms | Fever | 161 | 81.20 | 0.03-0.04 | **0.004** |
|  | Malaise | 17 | 8.63 | 0.06-0.27 |  |
|  | Weakness | 5 | 2.54 | 0.072-1.65 |  |
|  | Weight lost | 6 | 3.04 | 0.63-18.58 |  |
|  | Flu-like symptoms | 8 | 4.06 | 0.52-51.36 |  |
| Onset of symptoms | One day | 28 | 14.21 | 0.13-0.21 | 0.847 |
|  | 2-6 days | 161 | 81.72 | 0.62-0.87 |  |
|  | 7-13 days | 5 | 2.54 | 1.16-1.56 |  |
|  | >14 days | 3 | 1.52 | 0.68-4.55 |  |

### Risk factors associated with Brucella seropositivity

The association between the dichotomous outcome variable seroprevalence and potential risk factors was first examined in the univariable analysis (Table 7). All variables with p<0.25 were selected for further analysis to build the multivariable logistic regression model. In the multivariable logistic regression, only the number of animals (p<0.039) was statistically significantly associated with brucellosis (Table 8).

**Table 7:**
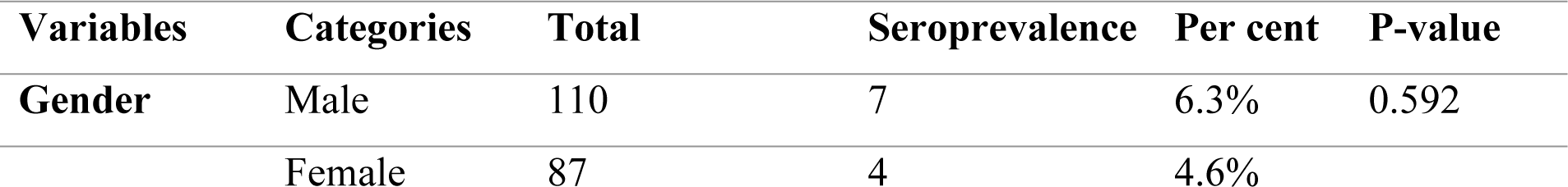
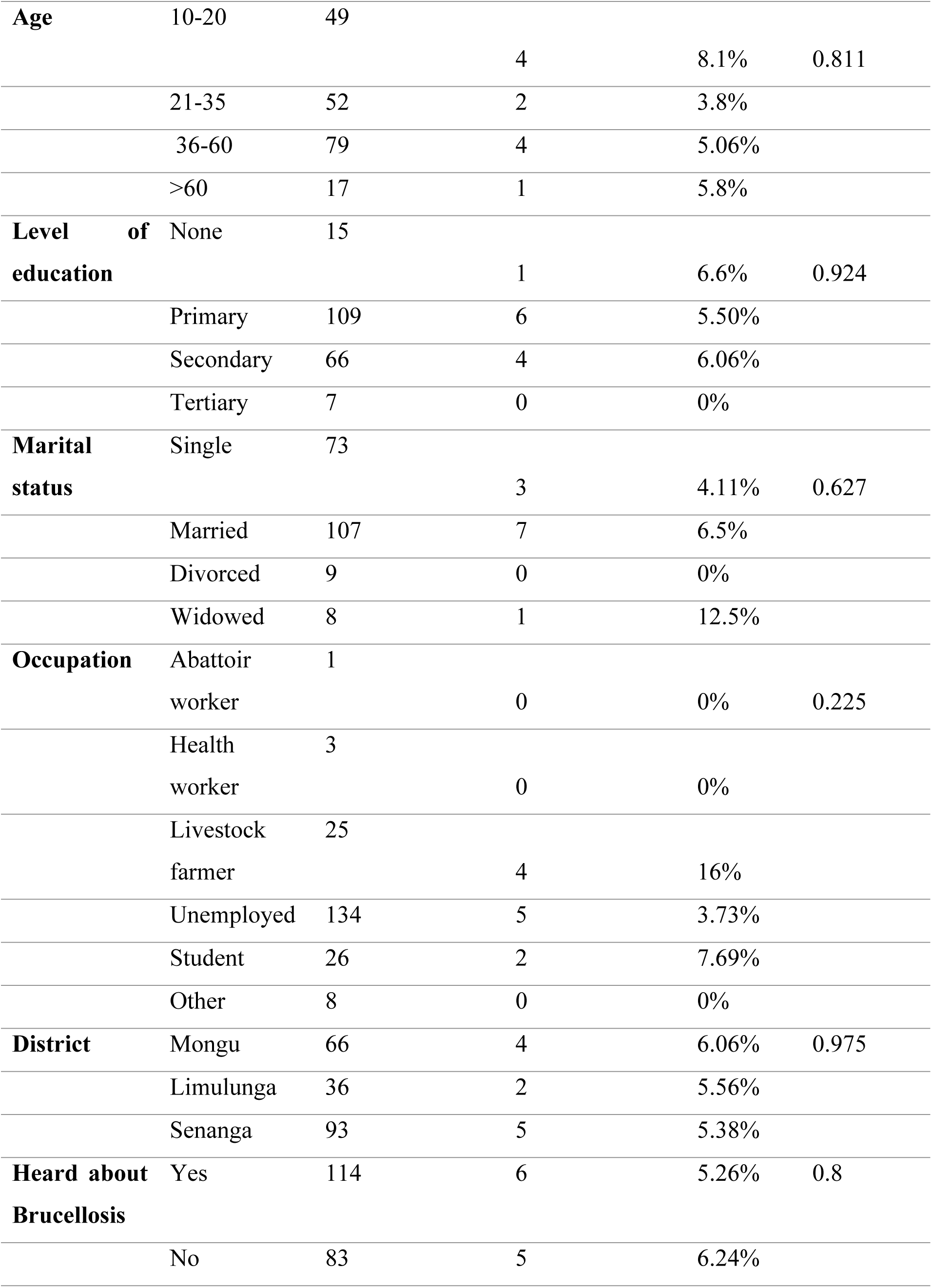
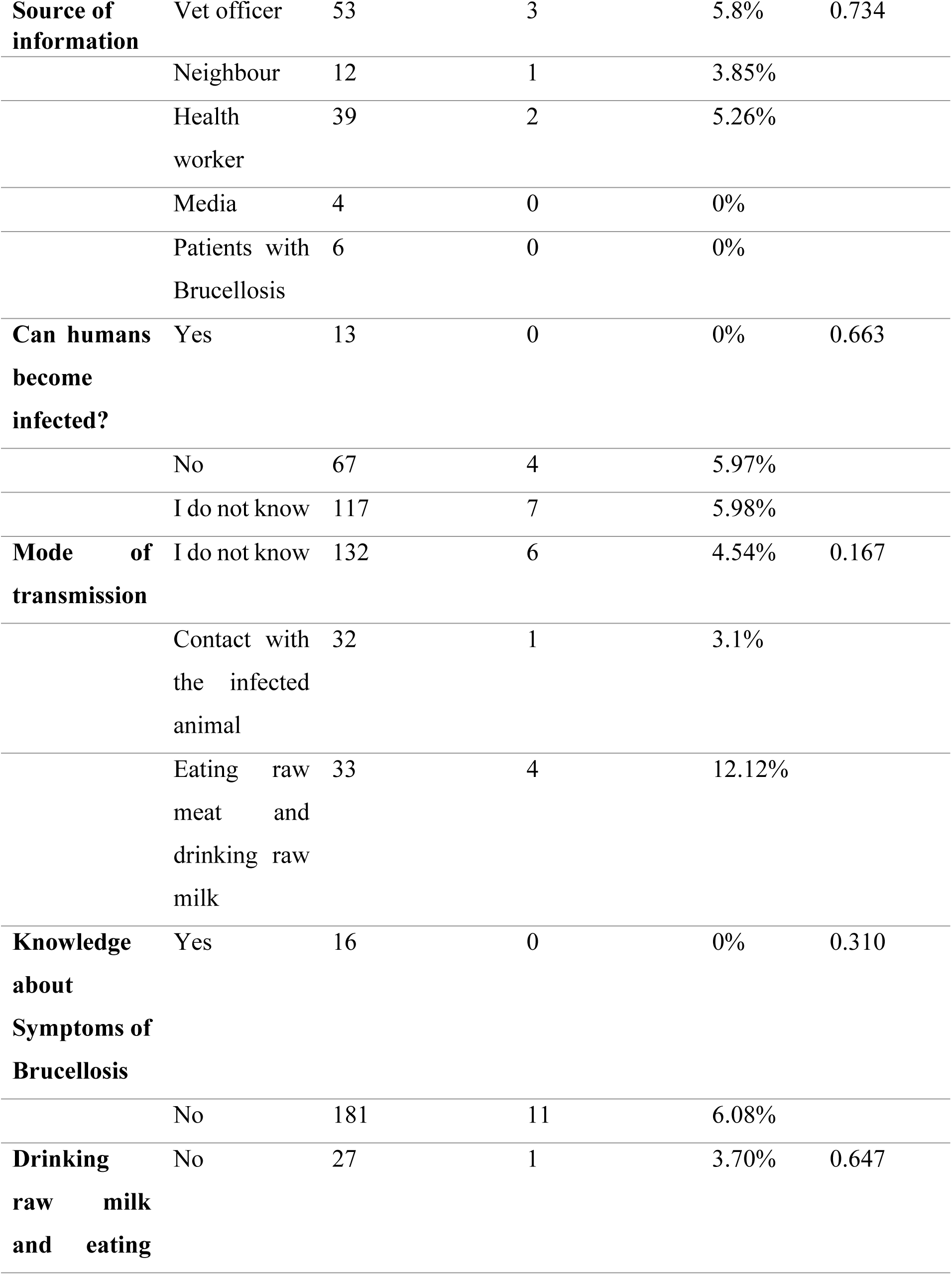
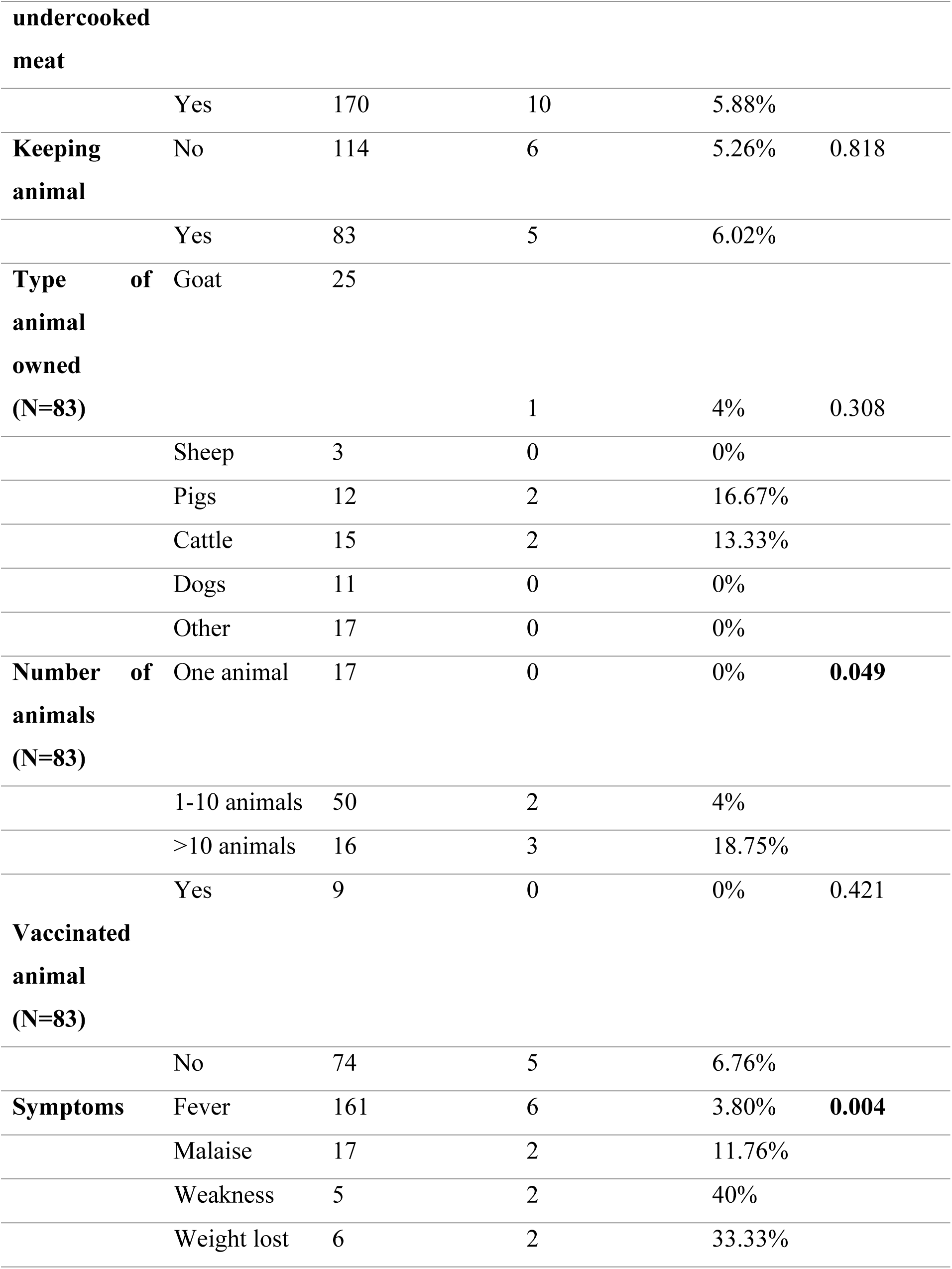
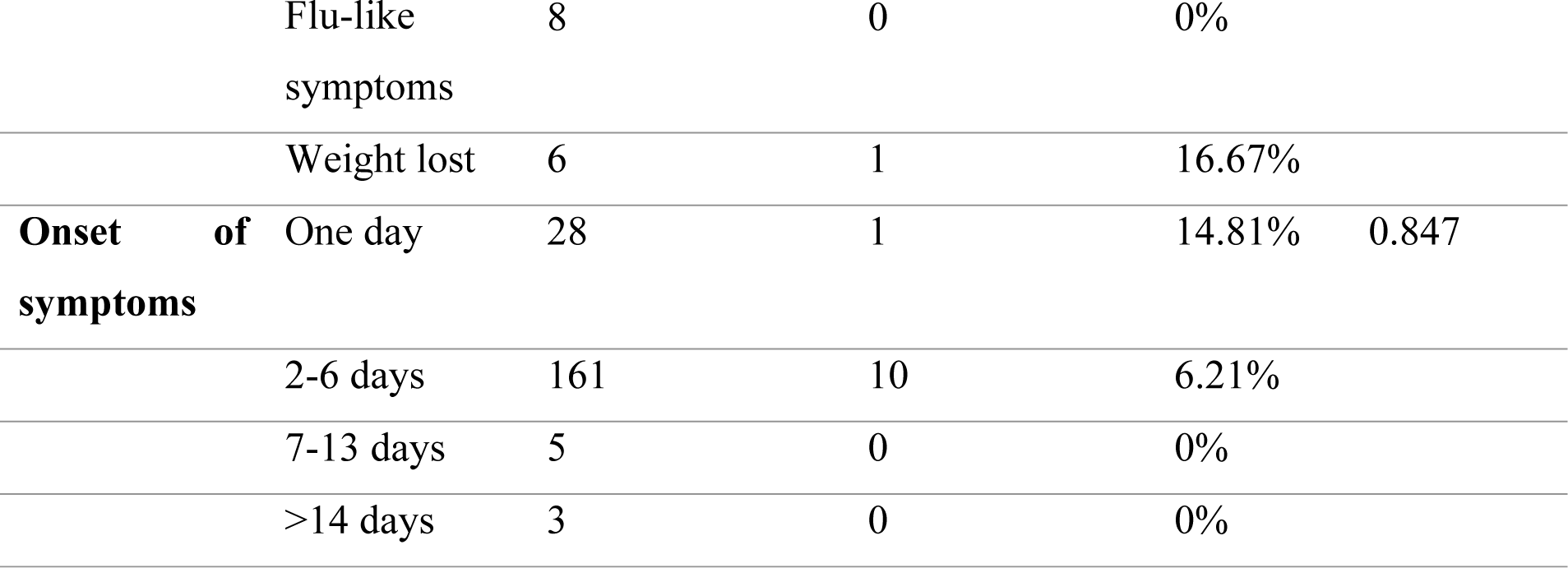
Univariable analysis of Potential Risk factors associated with Brucellosis.

**Table 8:** Standard Multivariable logistic regression analysis for brucellosis risk factors in humans.

| Variables | OR | (95%CI) | P-value |
| --- | --- | --- | --- |
| Type of animal owned | 0.72 | 0.42-1.23 | 0.226 |
| Number of animals | 6.49 | 1.10-38.13 | **0.039** |

## Discussion

To our knowledge, this is the first study to document the seroprevalence of *Brucella* antibodies among febrile patients seeking medical attention in community hospitals in the Western province of Zambia.

### Seroprevalence of human *Brucella* antibodies

The seroprevalence of human *Brucella* antibodies among patients seeking medical attention was 4.57%. The current study was based on community health facilities and highlights the seropositivity in patients from three districts (Mongu, Senanga and Limulunga) in the Western Province of Zambia. The 4.57% seroprevalence in our study almost corroborates the 5.03% earlier reported in Southern province^29^ and the 6% found among febrile patients attending a District hospital in Rwanda^30^.

However, the seroprevalence in this study is lower than the 20.2% reported in the Southern Province of Zambia^31^. While this present study was focused on screening febrile patients seeking medical attention, that of Mubanga et al targeted occupationally exposed humans (herdsmen and abattoir workers), who were at high risk of exposure to brucellosis ^31^. This could explain the observed differences in the studies.

Our study findings are also lower than the 14.9% observed among community hospital patients in Southwestern Uganda^32^. This variation can be due to various possible reasons. Southwestern Uganda has a high rate of milk production, with a daily output of 100,000 litres, accounting for 35% of the country’s total production. As a result, the population in this area is likely to be exposed to health risks due to cultural practices involving the consumption of raw milk.^32^ A similar study in Saudi Arabia reported a seroprevalence of 12.8% among patients with fever^33^. The difference could be ue to the fact that *Brucella melitensis* has been reported to be the most prevalent pathogen causing human brucellosis in Saudi Arabia, ^34^, while Zambia has reported *Brucella abortus* ^34^. *Brucella melitensis* is known to be the most pathogenic species among the *Brucella* species ^35^.

### Knowledge and attitude of participants about Brucellosis

More than half of the respondents (57.87%) were knowledgeable about the disease. Some studies have demonstrated low knowledge levels of the disease as a risk factor that increases the risk of *Brucella* infection in the community, has a detrimental effect on *brucellosis* control measure compliance, and may contribute to underreporting of disease incidence in the nation ^36,37^. In contrast, only 6.60% of participants were aware that *Brucella* can affect humans, and most respondents 67.01% were ignorant about the mode of transmission to humans, which agrees with the study findings reported in Uganda ^38^ and Jordan ^39^. This is also similar to the findings by Munyeme et al. who noted that the population’s low awareness of *Brucellosis* was caused by a lack of health education programs, inadequate training in handling and rearing animals, a lack of extension services, the absence of health facilities, and remote participant locations^40^. The low awareness levels of human brucellosis in this study may be attributed to the limited formal education received by individuals in the research locations. Due to the lack of education on zoonotic diseases, farmers may not be well-informed about transmission paths, which could lead to a lack of preventative measures against brucellosis, as explained by Bouzoukeev^29^. *Brucella* seropositivity was higher in Senanga district than in Mongu and Limulunga districts. This can be attributed to the fact that most of the samples collected in Senanga were from rural health centres, while in Mongu and Limulunga districts, most samples were collected from mini hospitals. In rural health centres, poor knowledge levels concerning the disease can positively impact preventive measures regarding consuming raw meat and unpasteurised milk, which are major risk factors in the transmission of Brucellosis^42^. According to Ruano and Aguayo, low levels of awareness like those found in the current study put the population at risk of contracting *Brucella* and have a detrimental effect on *brucellosis* control measures compliance^31^. It may cause underreporting of disease occurrence in the nation. It is also noted that some rural people keep animals in their houses^43^. Access to personal medical or veterinary care to educate people about the disease is sometimes very difficult compared to urban areas. Moreover, most medical personnel have a low brucellosis suspicion rate among febrile patients ^44^. Similarly, a case-control study in Iran demonstrated awareness regarding modes of brucellosis transmission, for example consuming raw milk cheese was associated with a reduced risk of human brucellosis^43^.

### Risk factors associated with *Brucella* seropositivity

In this study, it was found that the number and types of animals were statistically significant risk factors that were associated with *Brucella* seropositivity. The close contact between domestic animals and human beings is still a critical mode of disease transmission^49,50^. Most humans have animals that can transmit *Brucella* pathogens, so the exposure risk is higher. Among the number of animals, those who had more than ten animals, 23.08% (3/13) were more likely to be exposed^46^. The importance of the number of animals favours migration or mobility, which increases the risk of Brucellosis transmission ^47^. The study conducted in Kenya by Kairu et al. found the number of animals was a significant risk factor^48^ with larger herds having a significantly higher risk of exposure to the disease. Large herds are often associated with poor sanitation, clustering of animals, and mixing of animals from different herds and species. Several studies in Africa have revealed a consistent correlation between brucellosis seropositivity and the number of animals ^48^. Another study in Mexico found that the number of animals is a risk factor^51^.

The sample size was less than the target but enough to carry out the planned analysis of this study without affecting its validity. However, some of the samples were hemolysed, and it is possible that other positive cases were not detected due to the exclusion of these samples. Despite these limitations, the study findings provide information on the seroprevalence of *Brucella* antibodies and associated risk factors among febrile patients in community hospitals in Western province of Zambia.

## Conclusions

*Brucella* antibodies were present at 4.57% among patients in community hospitals in Western Province. Most participants were unaware that *Brucella* can affect humans and were ignorant about the mode of disease transmission to humans. The number and types of animals were significantly associated with *Brucella* seropositivity.

## Data Availability

All relevant data are within the manuscript and its Supporting Information files.

## Acknowledgements

We acknowledge the participants who took part in this study. We are also grateful to the University of Zambia for providing all the references in this publication.

## Ethical statement

Ethical approval for the study protocol was obtained from Excellence in Research Ethics and Science (ERES) (Ref No. 2018-Dec-004). The authority to conduct the research was granted by the National Health Research Authority (NHRA), while permission to conduct the study at the health facilities was obtained from the Provincial Health Director in Western Province before the commencement of the study. Written informed consent was obtained from all the participants enrolled in the study. For participants below 18 years, written informed consent was obtained from their parents or legal guardians, followed by assent, to ensure they understood the study before agreeing to participate voluntarily. The participant consent outlined the purpose of the study, procedures, risk of minimal pain and discomfort at the injection site during blood sample collection, benefits, and the right to withdraw at any time. The site of needle puncture during blood collection and their right to withdraw at any given time during the study. All data for the study were restricted to the investigators and treated in confidence, no participant identifiers were used.

## Transparency declarations

All authors declare no conflict of interest.

